# Impact of Covid-19 Control Strategies on Health and GDP Growth Outcomes in 193 Sovereign Jurisdictions

**DOI:** 10.1101/2025.04.08.25325452

**Authors:** Matt Boyd, Michael G Baker, Amanda Kvalsvig, Nick Wilson

## Abstract

**Background:** The Covid-19 pandemic has caused approximately 27.3 million excess deaths globally as of June 2024. Despite growing research on pandemic response factors, the effectiveness of different strategic approaches to Covid-19 control remains insufficiently investigated.

**Aims:** To examine associations between Covid-19 pandemic control strategies (including stringent border restrictions) with age-standardized excess mortality and GDP per capita growth outcomes during 2020–2021.

**Methods:** Analysis of 193 sovereign jurisdictions with existing Global Burden of Disease Study data. Jurisdictions were classified by implementation of exclusion/elimination strategies reported in published literature, and the level of border restriction measures based on the Oxford Stringency Index. Multivariable analyses adjusted for island status, GDP per capita, and an index of government corruption. Excess mortality was cube root transformed and GDP per capita log transformed for regression analysis.

**Results:** Jurisdictions implementing explicit exclusion/elimination strategies showed the lowest age-standardized excess mortality (–2.1/100,000) compared to others (166.5/100,000). Island jurisdictions experienced lower mortality (64.8/100,000) than non-islands (194.3/100,000). Duration of border restrictions correlated with reduced excess mortality in islands (Pearson’s r = –0.624, p <0.001; β –0.004, island interaction –0.005, p <0.001), but not in non-islands. However, this effect weakened when controlling for government corruption in a subsample (lower corruption was associated with lowered mortality). No consistent significant relationships emerged between border measures and GDP growth, suggesting that stringent border restrictions in a pandemic may not significantly harm economies.

**Conclusion:** Exclusion/elimination strategies and related stringent border restrictions were associated with better health outcomes, particularly for islands. Effectiveness was likely partially mediated by governance quality. Future pandemic planning should consider both control strategy selection and implementation context, both of which are modifiable.

## Introduction

The Covid-19 pandemic has exacted an extraordinary toll on global health and economic systems. Current estimates indicate pandemic-related cumulative excess deaths of 27.3 million (95% uncertainty interval: 19.3 to 36.3) as of June 2024 [1]. The economic impact has been similarly profound, with the International Monetary Fund projecting total macroeconomic losses due to the pandemic at $13.8 trillion through 2024 [2].

Future pandemics are considered likely [3], with potential for catastrophic outcomes should engineered pathogens be involved [4]. This risk may be heightened by rapid developments in artificial intelligence that could facilitate the development of such engineered pathogens [5].

Given the world now has five years of experience with managing Covid-19 it is important to reflect on which factors and responses were associated with differential Covid-19 pandemic outcomes. For example, research has found associations with trust in government [6], social cohesion [7], and pandemic preparedness [8].

In this paper we focus on investigating the role of strategic approaches to Covid-19 control, notably exclusion/elimination strategies compared with mitigation (including tighter suppression) as described in established strategic frameworks [9]. A major reason for focussing on strategies is that they are concerned with goals, rather than specific interventions such as the use of ‘lockdowns’. When a lockdown is used for elimination it is acting as a ‘circuit breaker’ for a few weeks until there is evidence of zero transmission of Covid-19 in a jurisdiction, and such measures can then be relaxed without fear of an immediate resurgence. Lockdowns used for mitigation have a different goal of ‘flattening the curve’ so they generally need to be used with varying intensity for sustained periods, which is different from their use for elimination.

Several studies have examined the relationship between country-level strategic responses to the Covid-19 pandemic and their subsequent health and economic outcomes. A study published in December 2020 documented more favourable health outcomes and GDP projections among countries pursuing elimination strategies (China, Taiwan, Australia, and New Zealand) compared to European and North American nations implementing mitigation and suppression approaches [9]. Similarly, research analysing 44 countries in 2021 found that five “elimination strategy countries” (Australia, China, Japan, New Zealand, and South Korea) experienced lower Covid-19 mortality rates and less severe effects on GDP growth than “suppression/mitigation strategy countries” [10].

Additional research published in 2021 corroborated these findings, reporting lower Covid-19 mortality rates among OECD countries utilising elimination strategies (Australia, Iceland, Japan, New Zealand, and South Korea) versus those implementing mitigation strategies [11]. This study further concluded that “elimination is superior to mitigation for GDP growth on average and at almost all time periods” [11].

A 2023 study constructed a Covid-19 pandemic “shock index” incorporating health, behavioural, and economic indicators for 44 countries [12]. It determined that compared to “reactive” countries, “proactive” nations (Australia, New Zealand, and South Korea) “consistently outperform the others both in terms of robustness and resilience,” further noting that “proactive countries better preserve their performance over time whereas similarly performing reactive ones slide down.”

Beyond economic and mortality metrics, mental health impacts have also been examined. A study of 15 countries with regular mental health surveys during 2020 and 2021 reported that “elimination strategies minimized transmission and deaths, while restricting mental health effects” [13]. The elimination strategy countries in this analysis included Australia, Japan, Singapore, and South Korea, though the study acknowledged that “changes in mental health measures during the first 15 months of the COVID-19 pandemic were small.”

Research has also examined intra-country perspectives on elimination strategies. Canadian studies reported that “the Atlantic provinces (New Brunswick, Nova Scotia, Prince Edward Island, and Newfoundland and Labrador) and Northern Canada (Nunavut, Yukon and Northwest Territories) generally implemented a containment strategy” consistent with an “elimination” or “zero-COVID strategy” [14]. This approach demonstrated some success, with these regions reporting “long periods with no community cases” [15]. Martignoni et al. concluded that “While elimination may be a preferable strategy for regions with limited healthcare capacity, low travel volumes, and few ports of entry, mitigation may be more feasible in large urban areas with dense infrastructure, strong economies, and with high connectivity to other regions.”

Border closures may be the most critical component of the elimination strategy, where it needs to be sustained for the duration of the response [16]. Hence it is useful to refer to this strategy as “exclusion/elimination”. If implemented proactively early in a pandemic, stringent border restriction has potential to fully exclude a pandemic agent [17].

At the onset of the global Covid-19 pandemic, most Pacific Islands and Territories “implemented rapid border closures” [18] which can be considered an “exclusion strategy”. These responses may have been generally effective given that one analysis has reported that Oceania had the lowest excess mortality rate of any region (for January 2020 to September 2022) [19]. Another analysis [20], found that Oceania (which included Australia in this work) had negative excess mortality in 2020 and a low positive excess in 2021.

Another study of excess mortality during the Covid-19 pandemic reported that this was lower for island nations [7]. Particular nations that were considered to perform relatively well include: Australia, Iceland, Singapore, New Zealand and Taiwan [21, 22]. Being an island nation also significantly increased the probability of a country pursuing an exclusion/elimination strategy [10].

But in terms of quantifying the impact of border closure, there have been only a few studies. One study “found no evidence in favor of international border closures” while finding “a strong association between national-level lockdowns and a reduced spread of SARS-CoV-2 cases” [23]. While an analysis of 166 countries found that “total border closures banning non-essential travel from all countries and (to a lesser extent) targeted border closures banning travel from specific countries had some effect on temporarily slowing COVID-19 transmission in those countries that implemented them” [24].

### Aims and hypotheses

In light of the background above, we aimed to explore any associations between Covid-19 control strategies with both excess mortality and GDP growth outcomes. Specific hypotheses were as follows:

- Hypothesis One (strategic choices) was that the five jurisdictions that clearly used an exclusion/elimination strategy would have lower age-standardized excess mortality than other jurisdictions. We refer to this group as having ‘explicit’ exclusion/elimination strategies.
- Hypothesis Two (implementation of control measures) was that the jurisdictions using relatively fast and complete border closure (based on date of border closure and stringency of border restrictions) would have lower age-standardized excess mortality than other jurisdictions (when adjusted for GDP per capita and island status). We refer to this group as having stringent border controls.
- Hypothesis Three was that both the above hypotheses would also apply to a relatively successful macroeconomic response in terms of GDP growth.

## Materials and methods

Ethical review not required for this analysis of jurisdiction-level data.

### Study jurisdictions

The study comprised 193 sovereign jurisdictions with excess mortality data for 2020– 2021 from the Global Burden of Disease (GBD) study [25]. We used the super-region and region classifications from the GBD. Additional categorisation was performed for island (n=48) vs non-island status (n=145). In the main analyses we included as islands those jurisdictions that had a land border with another jurisdiction on the same island (eg, Dominican Republic/Haiti, Ireland/UK, Papua New Guinea/Indonesia, and Timor Leste/Indonesia etc). We defined islands as per previous work [26].

We excluded non-sovereign island states because it is not always clear what role the local government played in pandemic-related decision-making (as opposed to the colonial power). Namely: American Samoa, Bermuda, Cook Islands, Greenland, Guam, Niue, Northern Mariana Islands, Puerto Rico, Tokelau, Virgin Islands. We also excluded non-sovereign non-island jurisdictions (eg, Hong Kong). We excluded North Korea as data seemed unreliable with conflicting reports.

### Identifying explicit exclusion/elimination strategy jurisdictions

As detailed in the *Introduction*, all of the following jurisdictions have been described in at least one study as adopting an elimination strategy: Australia, China, Iceland, Japan, New Zealand, Singapore, South Korea, and Taiwan. But based on our own work relating to Iceland [27], we consider “mitigation/suppression” is more appropriate for that country. Also Japanese authors have explained why Japan did not use an elimination strategy [28].

Similarly, the South Korean Government never appeared to articulate an “elimination” or “zero-Covid” goal, and so we think a “mitigation/suppression” strategy is the most accurate description (albeit at a very high level of control during 2020). Therefore, our analysis defined just five ‘explicit’ exclusion/elimination strategy jurisdictions: Australia, China, New Zealand, Singapore, and Taiwan.

### Identifying jurisdictions that used stringent border restrictions

For classifying border restrictions we considered the Oxford Covid-19 Government Response Tracker [29], specifically the date at which jurisdictions were coded to have reached border control stringency ‘4’, meaning a ban on arrivals from all regions or total border closure (note this index does not report information pertaining to citizens of the jurisdiction who may have required levels of home or facility-based quarantine). We obtained this stringency data as published by Our World in Data [30], and filled data gaps where possible, expanding the dataset by eight additional jurisdictions based on analyst and media reporting and/or government documents. We specifically determined:

- Time until closure (days from 1 January 2020 until level ‘4’ implemented)
- Duration of closure (sum of days spent at level ‘4’)
- Time until relaxing closure (days from 1 January 2020 until last level ‘4’ day)

### Outcome data sources

#### Excess mortality data

Excess mortality and age-standardized excess mortality by year, for all the jurisdictions (2020 and 2021) was obtained from the GBD Study Demographics Collaborators (see Acknowledgements). The estimation methods for these data have previously been described in a GBD publication [25]. We consider that excess mortality is an important measure of the true mortality impact from the Covid-19 pandemic given that it identifies the difference between observed all-cause mortality and mortality expected under normal conditions. Consequently, this measure is not subject to the high levels of under ascertainment seen with Covid-19 mortality surveillance [31].

#### GDP per capita and GDP growth

Given that excess mortality in 2020–2021 is associated with level of development (eg, a sociodemographic index [32]), we treated GDP per capita as a covariate in the multivariable analyses. GDP data for these jurisdictions was sourced from the World Bank’s World Development Indicators, namely GDP per capita purchasing power parity (PPP) (constant 2017 international $), which included adjustment for inflation, ie, the World Bank dataset “NY.GDP.PCAP.PP.KD”, and calculated GDP growth from 2019 to 2020, and from 2020 to 2021, based on five-year geometric means.

#### Control variables

Our primary control variable was mean GDP per capita (as above) for the five-year period ending in 2019. We also obtained data from the Covid-19 NP Collaborators for the three variables they found consistently associated with Covid-19 outcomes [6]. These data represented the level of government corruption, trust in individuals, and trust in government. The data had been developed through principal components analysis of underlying global survey data, as described by the Covid-19 NP Collaborators in 2022 [6]. However, with the exception of government corruption, data was missing for the majority of island jurisdictions limiting potential inclusion.

#### Statistical analysis

We established Pearson’s r correlations across relationships and performed multivariable linear regressions controlling for island status, GDP per capita and government corruption. To examine border restriction patterns and their associations with key outcomes, we categorized jurisdictions based on their implementation of level 4 border restrictions.

After excluding jurisdictions with missing data, we created three groups: those that never reached level 4 border restrictions (0 days), those with a duration below the median number of days at level 4, and those above the median. We compared age-standardized excess mortality, GDP growth 2019-2020, and GDP growth 2020-2021 across these groups using one-way ANOVA. Mean values with standard deviations were calculated for each group. To account for geographical and jurisdictional characteristics, we conducted stratified analyses for islands, non-island jurisdictions, and specific regions (Oceania and Caribbean). For each stratum, we reported sample sizes, means with standard deviations, and p-values from ANOVA tests comparing the three level 4 restriction groups.

## Results

### Global analysis of excess mortality by GBD region and explicit exclusion/elimination status

Our analysis of 193 jurisdictions revealed substantial variation in Covid-19 mortality outcomes and control strategies across different geographic and socioeconomic contexts (Table 1). The overall age-standardized excess mortality for the 2020 to 2021 period was 162.1 per 100,000 population (SD=154.4), with non-island jurisdictions experiencing significantly higher excess mortality rates (194.3) compared to island jurisdictions (64.8). Notably, explicit exclusion/elimination jurisdictions (n=5), demonstrated negative excess mortality (–2.1), in stark contrast to non-exclusion/elimination jurisdictions (166.5) (Table 1, Fig 1).

**Fig 1.**
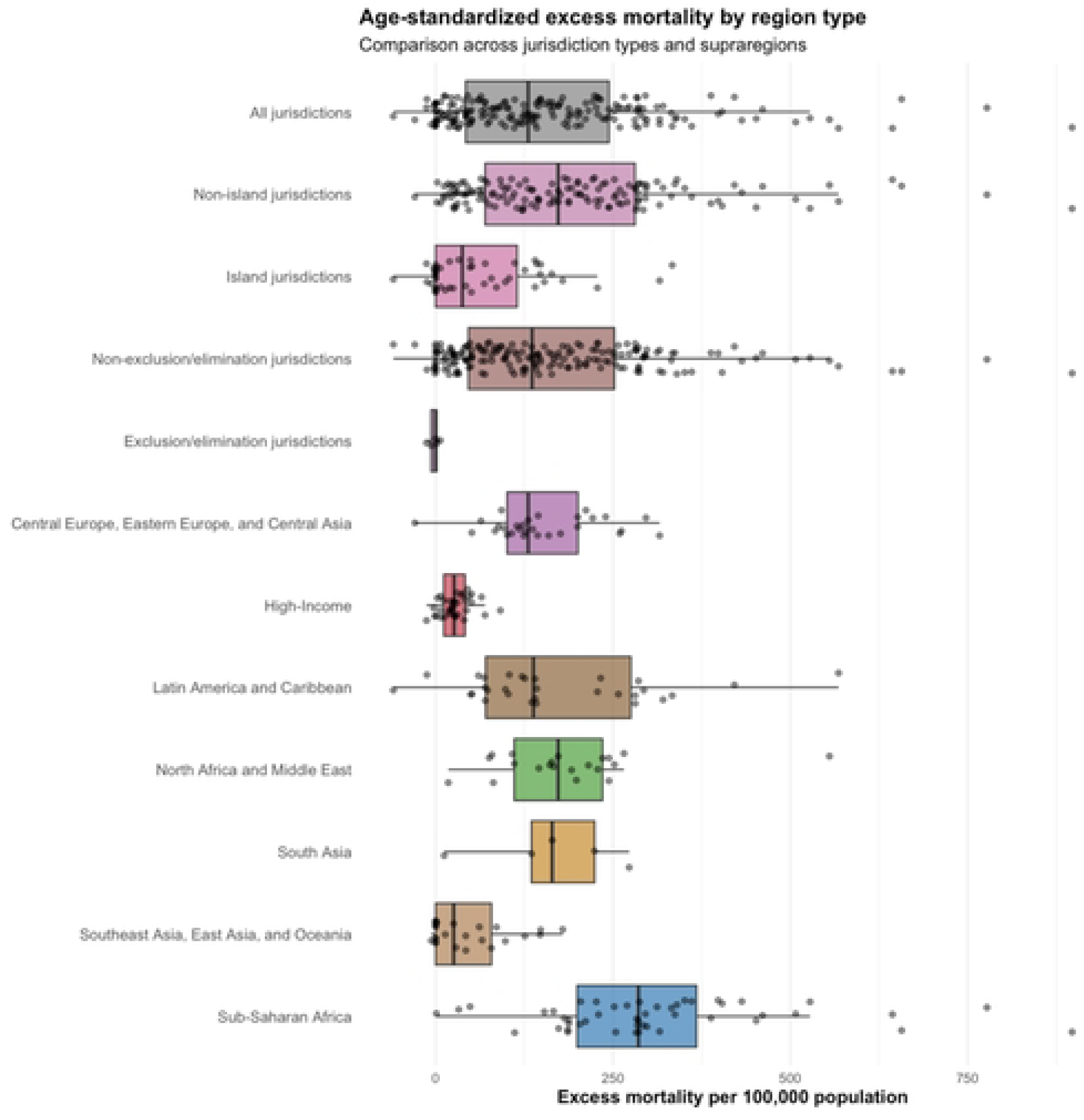
Age-standardized excess mortality (2020–2021). By jurisdiction, exclusion/elimination strategy, and GBD super-region, with box plot of median value, upper and lower quartiles, and range (note ‘high income’ region consists of several high-income countries across different parts of the world).

**Table 1:** Descriptive statistics by jurisdiction, exclusion/elimination strategy, and GBD region for the 2020 to 2021 period (note ‘high income’ region consists of several high-income countries across different parts of the world, ‘level 4’ is the maximum Oxford Stringency Index border restriction)

| Group | N | Age-standardized excess mortality (SD) | Number reaching level 4 border control (%) | Median days at level 4 (IQR) |
| --- | --- | --- | --- | --- |
| **Jurisdiction types** |  |  |  |  |
| All jurisdictions | 193 | 162.1 (154.4) | 159 (82.8%) | 130.5 (59-224) |
| Non-island jurisdictions | 145 | 194.3 (158.9) | 120 (83.3%) | 123.5 (59-207) |
| Island jurisdictions | 48 | 64.8 (84.8) | 39 (81.2%) | 160.5 (66.2-418) |
| Non-exclusion/elimination<br>jurisdictions | 188 | 166.5 (154.1) | 156 (83.4%) | 128.0 (60-221.5) |
| Explicit<br>exclusion/elimination<br>jurisdictions (see text) | 5 | -2.1 (7.5) | 3 (60.0%) | 200.0 (0-591) |
| **Super-region** |  |  |  |  |
| Central Europe, Eastern<br>Europe, and Central Asia | 29 | 150.4 (77.7) | 25 (86.2%) | 86.0 (56-137) |
| High-Income | 35 | 27.8 (22.1) | 18 (51.4%) | 59.0 (0-268) |
| Latin America and Caribbean | 30 | 170.9 (135.0) | 25 (83.3%) | 143.0 (89.2-199) |
| North Africa and Middle<br>East | 21 | 186.4 (108.9) | 18 (85.7%) | 138.0 (89-238) |
| South Asia | 5 | 161.7 (99.2) | 5 (100.0%) | 137.0 (61-163) |
| Southeast Asia, East Asia,<br>and Oceania | 25 | 45.5 (56.4) | 22 (88.0%) | 348.0 (143-693) |
| Sub-Saharan Africa | 48 | 311.7 (175.5) | 46 (97.9%) | 142.0 (106.5-208) |
| **Region** |  |  |  |  |
| Andean Latin America | 3 | 423.5 (143.6) | 3 (100.0%) | 199.0 (166-243.5) |
| Australasia | 2 | -6.4 (9.1) | 2 (100.0%) | 650.0 (620.5-679.5) |
| Caribbean | 16 | 114.0 (102.0) | 13 (81.2%) | 109.0 (66.2-183.2) |
| Central Asia | 9 | 175.2 (103.0) | 8 (88.9%) | 143.0 (124-406) |
| Central Europe | 13 | 140.5 (69.4) | 11 (84.6%) | 70.0 (21-84) |
| Central Latin America | 9 | 196.7 (99.4) | 7 (77.8%) | 153.0 (137-191) |
| Central Sub-Saharan Africa | 6 | 334.2 (89.0) | 5 (100.0%) | 144.0 (110-156) |
| East Asia | 2 | -2.2 (5.6) | 1 (50.0%) | 100.0 (50-150) |
| Eastern Europe | 7 | 136.9 (55.3) | 6 (85.7%) | 107.0 (57.5-119) |
| Eastern Sub-Saharan Africa | 17 | 286.2 (161.1) | 16 (94.1%) | 121.0 (36-168) |
| High-Income Asia Pacific | 4 | 7.7 (9.1) | 2 (50.0%) | 188.0 (0-410) |
| High-Income North America | 2 | 69.0 (31.5) | 1 (50.0%) | 254.5 (127.2-381.8) |
| North Africa and Middle East | 21 | 186.4 (108.9) | 18 (85.7%) | 138.0 (89-238) |
| Oceania | 12 | 34.4 (65.4) | 12 (100.0%) | 768.0 (359.8-854) |
| South Asia | 5 | 161.7 (99.2) | 5 (100.0%) | 137.0 (61-163) |
| Southeast Asia | 11 | 66.4 (42.9) | 9 (81.8%) | 145.0 (83.5-278.5) |
| Southern Latin America | 3 | 53.5 (23.9) | 3 (100.0%) | 385.0 (373.5-394) |
| Southern Sub-Saharan Africa | 6 | 601.1 (205.2) | 6 (100.0%) | 216.0 (207.8-227.2) |
| Tropical Latin America | 2 | 132.0 (14.2) | 2 (100.0%) | 164.5 (144.2-184.8) |
| Western Europe | 24 | 27.4 (14.5) | 10 (41.7%) | 0.0 (0-87.5) |
| Western Sub-Saharan Africa | 19 | 236.1 (93.5) | 19 (100.0%) | 144.0 (114-202.5) |

Values are presented as mean (SD) for mortality, count (%) for level 4 incidence, and median (IQR) for days at level 4.

Geographically, Sub-Saharan Africa exhibited the highest excess mortality among super-regions (311.7), while High-Income regions experienced substantially lower rates (27.8). Among the GBD regions, Southern Sub-Saharan Africa reported the highest mortality (601.1), whereas East Asia showed the lowest (–2.2).

The vast majority of jurisdictions (82.8%) reached level 4 border restrictions, with a median duration of 130.5 days (IQR: 59-224). Western Europe stood out for having the lowest proportion of jurisdictions reaching level 4 restrictions (41.7%), while Oceania jurisdictions maintained the longest median duration at this level (768 days), highlighting diverse policy approaches and pandemic trajectories across regions.

### Global analysis of outcomes based on border restriction measures

Among jurisdictions that reached level 4 border restrictions, calculation of Pearson’s r revealed no correlation between the number of days to enact level 4 and outcomes. However, there were strong negative correlations, particularly in island jurisdictions, between age-standardized excess mortality and duration of level 4 restrictions (p < 0.001), see Table 2.

**Table 2:** Correlations between border restrictions and outcomes by jurisdiction type (only for the 159 (83%) of jurisdictions that enacted the highest level of border restrictions ie, Oxford Stringency Index level ‘4’)

| Measure | Outcome | All jurisdictions | Islands | Non-islands |
| --- | --- | --- | --- | --- |
| **Age-standardized excess mortality** |  |  |  |  |
| Days until level 4 | Age-standardized excess mortality | 0.031 (n=159) | -0.256 (n=39) | 0.112 (n=120) |
| Days at level 4 | Age-standardized excess mortality | -0.473*** (n=159) | -0.624*** (n=39) | -0.127 (n=120) |
| Days until relaxation | Age-standardized excess mortality | -0.363*** (n=159) | -0.639*** (n=39) | -0.031 (n=120) |
| **GDP growth 2019-2020** |  |  |  |  |
| Days until level 4 | GDP growth 2019-2020 | 0.049 (n=151) | -0.090 (n=36) | 0.078 (n=115) |
| Days at level 4 | GDP growth 2019-2020 | 0.056 (n=151) | 0.294 (n=36) | -0.069 (n=115) |
| Days until relaxation | GDP growth 2019-2020 | 0.023 (n=151) | 0.262 (n=36) | -0.077 (n=115) |
| **GDP growth 2020-2021** |  |  |  |  |
| Days until level 4 | GDP growth 2020-2021 | -0.101 (n=150) | -0.072 (n=36) | -0.114 (n=114) |
| Days at level 4 | GDP growth 2020-2021 | -0.054 (n=150) | 0.137 (n=36) | -0.102 (n=114) |
| Days until relaxation | GDP growth 2020-2021 | -0.116 (n=150) | 0.100 (n=36) | -0.173 (n=114) |

\* $p < 0.05$ , \*\* $p < 0.01$ , \*\*\* $p < 0.001$ ; Values shown are Pearson correlation coefficients with significance levels asterisked and sample sizes (n)

Indeed, in this group the island interaction was highly statistically significant, when controlling for GDP per capita, and more than doubled the total effect of full border closure stringency on age-standardized excess mortality for islands (Table 3). Fig 2 illustrates the regression results for islands versus non-islands.

**Fig 2.**
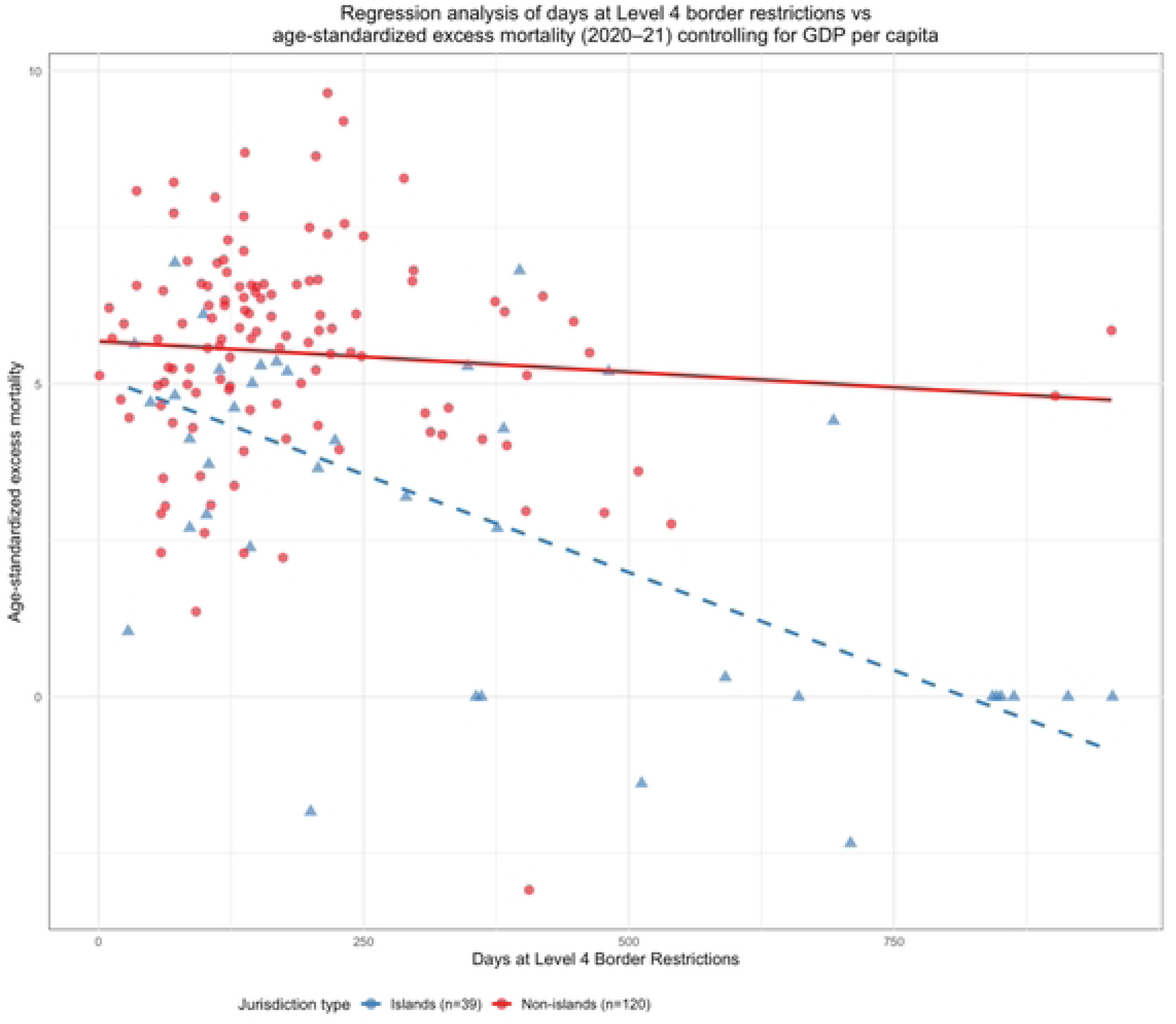
Regression results for jurisdictions reaching Level 4 border restrictions. Showing duration of restrictions vs excess mortality (2020–2021) outcome for non-islands (red) and island jurisdictions (blue).

**Table 3:** Regression analysis of jurisdictions reaching level 4 border restrictions and outcomes (excess mortality and GDP growth)

| Measure | Outcome | Main effect | Island<br>interaction | N |
| --- | --- | --- | --- | --- |
| **Age-standardized excess mortality** |  |  |  |  |
| Days until level 4 | Age-standardized EM | -0.002 | -0.012 | 151 |
| Days at level 4 | Age-standardized EM | -0.004*** | -0.005*** | 151 |
| Days until relaxation | Age-standardized EM | -0.003*** | -0.005*** | 151 |
| **GDP growth 2019-2020** |  |  |  |  |
| Days until level 4 | GDP growth 2019-2020 | 0.001 | -0.006 | 151 |
| Days at level 4 | GDP growth 2019-2020 | 0.001 | 0.004 | 151 |
| Days until relaxation | GDP growth 2019-2020 | 0.000 | 0.003 | 151 |
| **GDP growth 2020-2021** |  |  |  |  |
| Days until level 4 | GDP growth 2020-2021 | -0.004 | -0.000 | 150 |
| Days at level 4 | GDP growth 2020-2021 | 0.000 | 0.004 | 150 |
| Days until relaxation | GDP growth 2020-2021 | -0.001 | 0.003 | 150 |

\* $p < 0.05$ , \*\* $p < 0.01$ , \*\*\* $p < 0.001$ ; EM = excess mortality; Main effect shows coefficient for border measure controlling for GDP and island status. Island interaction shows additional effect for island jurisdictions. N is the number of observations used in each regression model.

However, results of one-way ANOVA analyses stratified by jurisdiction type, region, and GDP per capita (tertiles), revealed a more mixed picture (Table 4). Jurisdictions that never implemented level 4 border restrictions often showed the lowest age-standardized excess mortality rates. For countries in the highest GDP tertile—especially non-island nations—GDP growth was more favorable in places that had “below median” duration of level 4 restrictions compared to those with either “above median” duration or no level 4 restrictions at all. Interestingly, Oceania showed a different pattern. There, GDP growth was actually higher in jurisdictions with “above median” duration of level 4 restrictions compared to those with “below median” duration (2.6% vs –1.1% for 2019-20, p = 0.004; and 1.2% vs – 2.1% for 2020-21, p = 0.049). It’s worth noting, however, that in Oceania, we didn’t have any jurisdictions that completely avoided level 4 border restrictions for comparison.

**Table 4:**
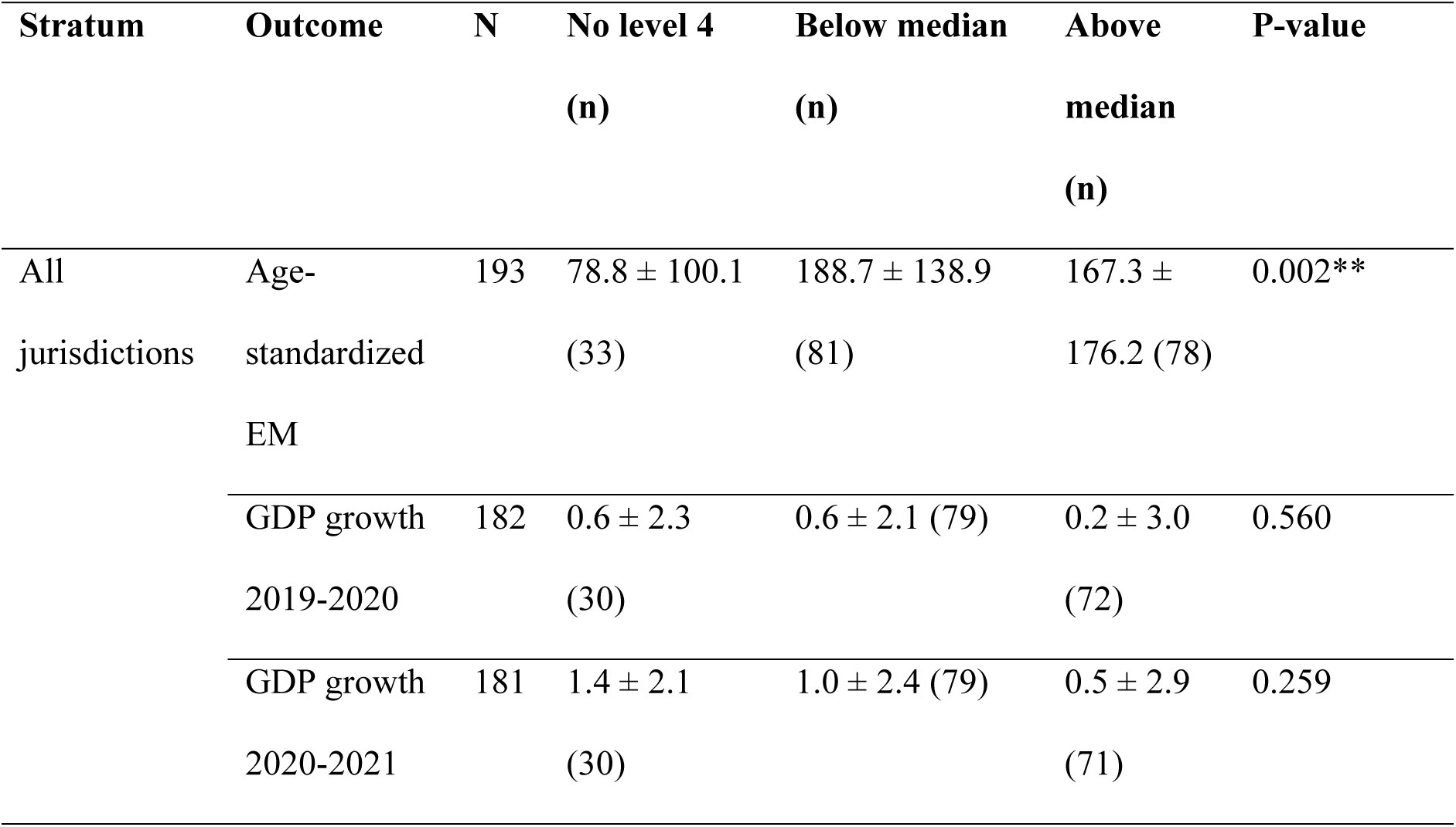
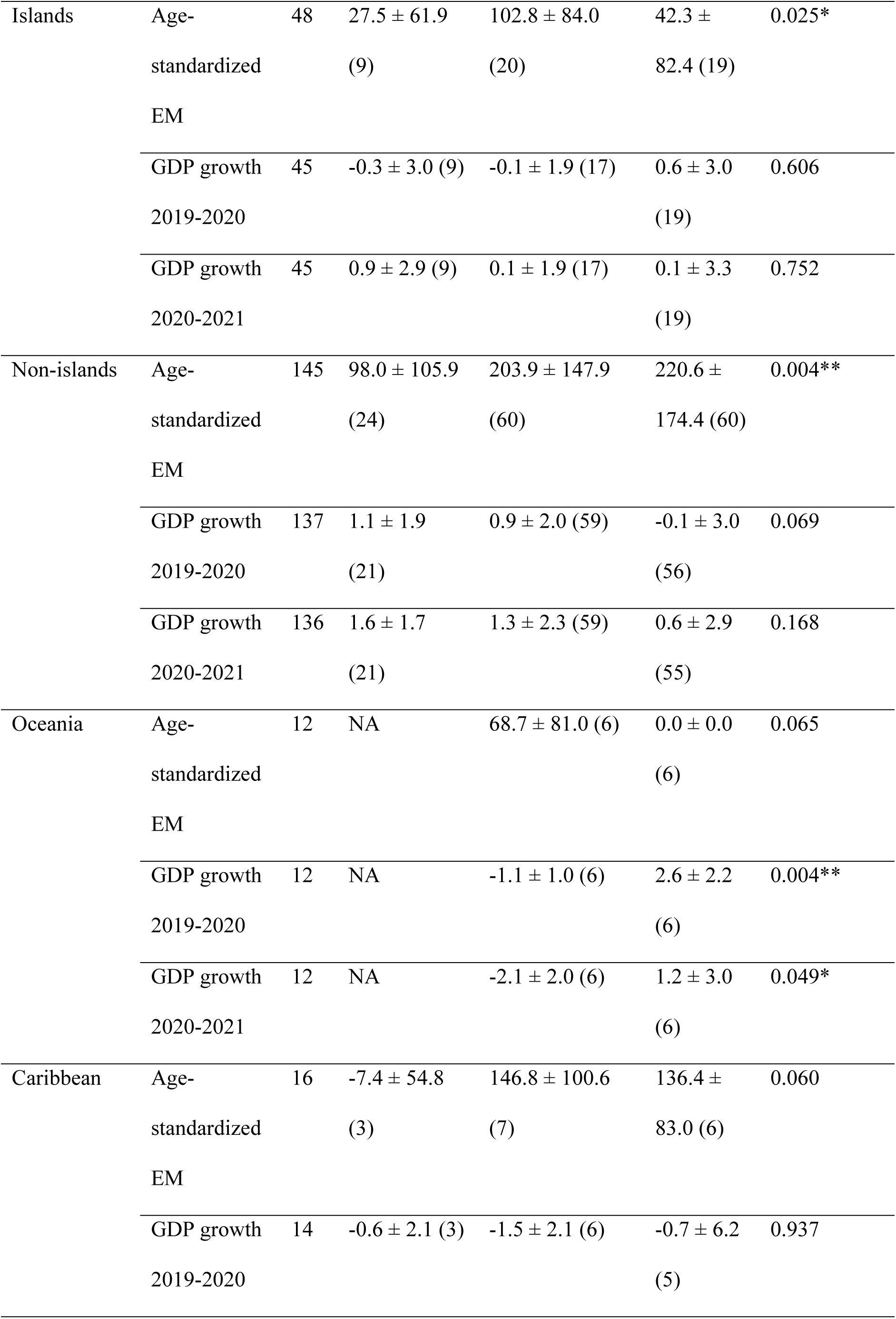
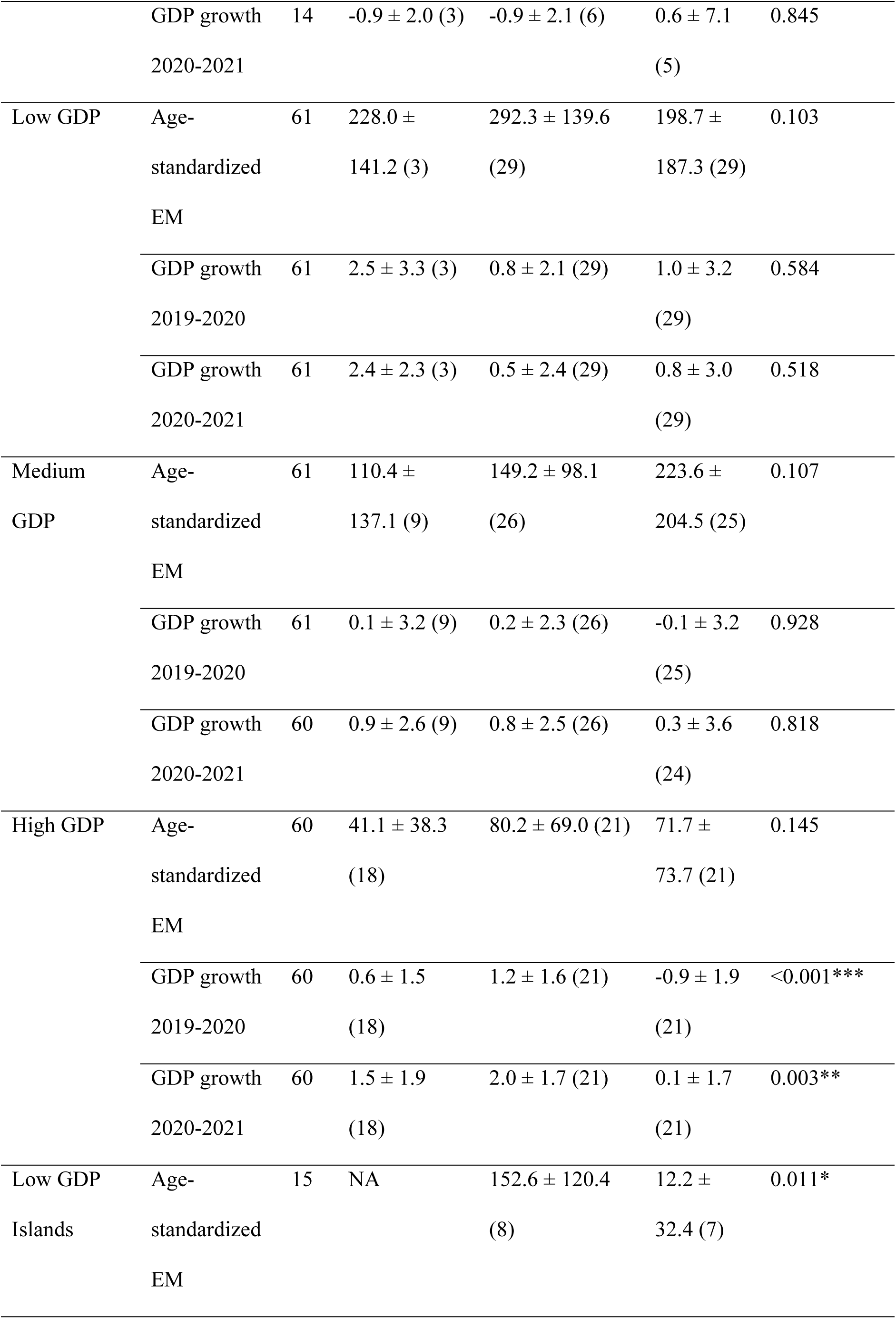
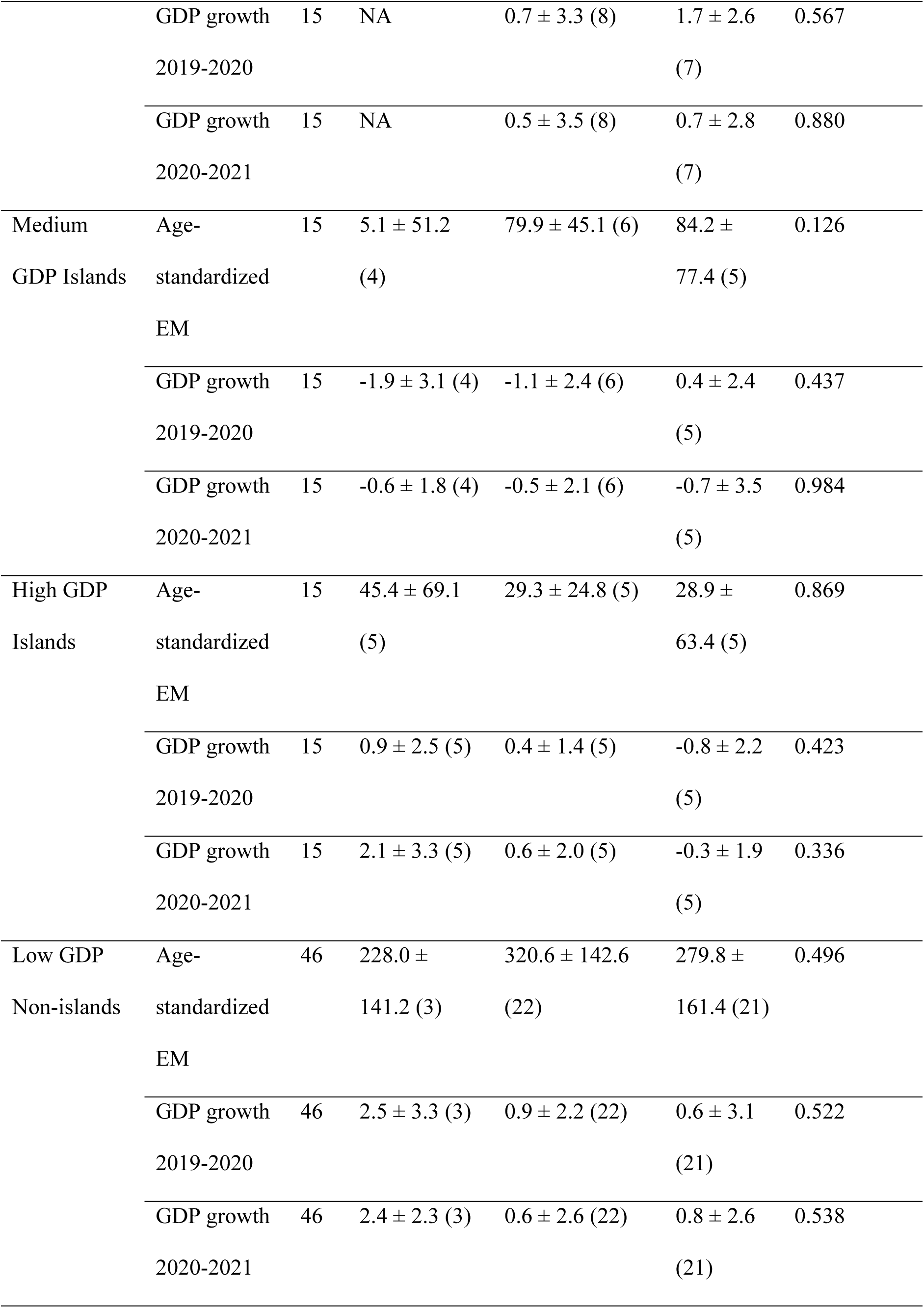
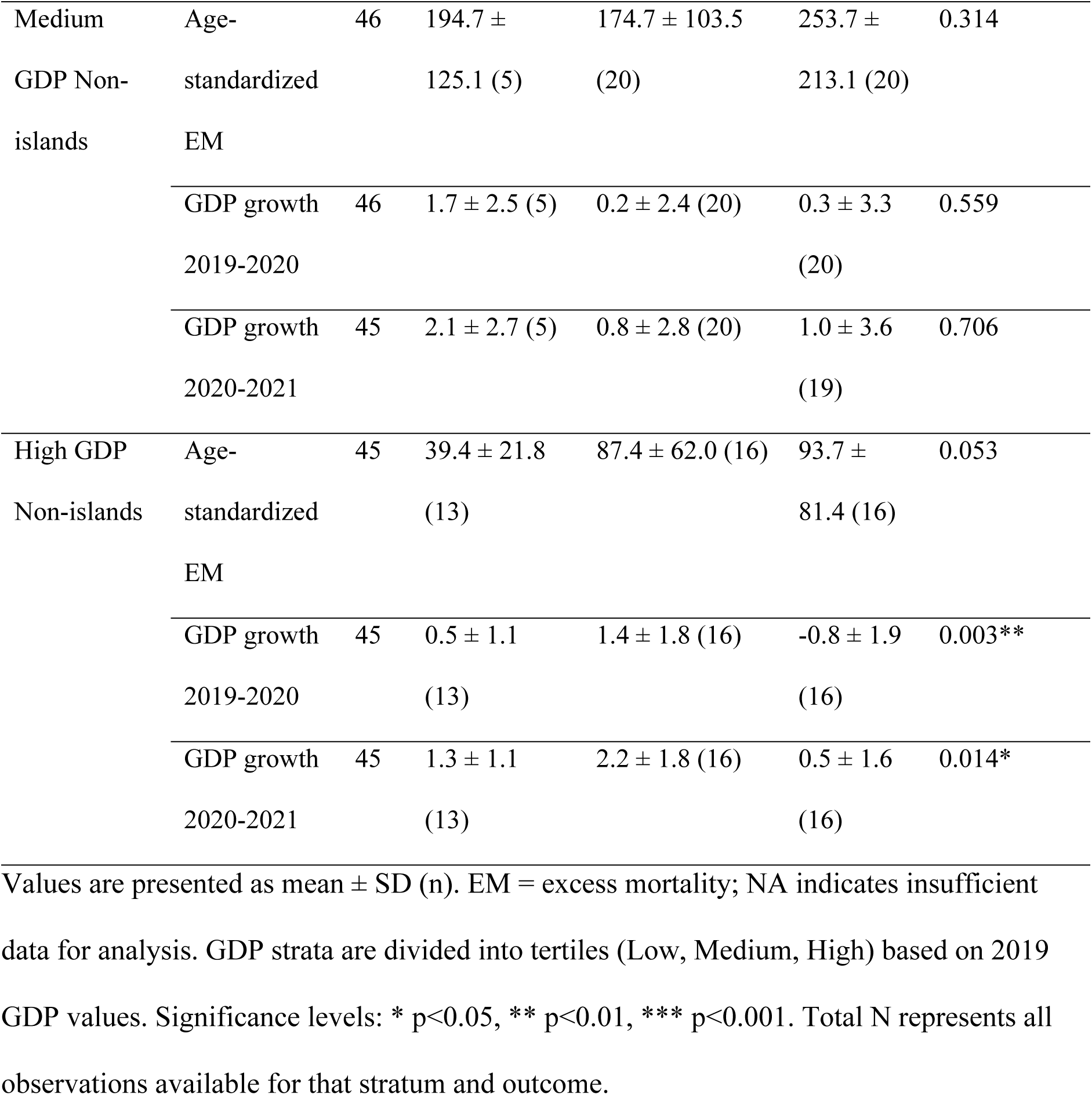
Results of one-way ANOVA comparing border closure stringency category (ie, no ‘level 4’ closure, below median duration, and above median duration of ‘level 4’ within each stratum) when stratified by island status, island region, and GDP per capita (tertiles).

### Analysis of outcomes restricted to island jurisdictions

When we took islands (n = 36) forward for regression analysis in isolation (based on the findings in Table 2 and Table 3 that demonstrated strong correlation between excess mortality outcomes and duration of border measures for islands enacting Level 4 restrictions, but not for non-island jurisdictions), we found that the duration of level ‘4’ border restrictions was strongly associated with reduced age-standardized excess mortality in island jurisdictions (Table 5). In our most significant model (R² = 0.576, F = 22.42, p < 0.001), each additional day at level 4 restrictions was associated with a reduction in the cube root of age-standardized excess mortality (β = –0.006, p < 0.001), while controlling for log-transformed GDP per capita (β = –0.902, p < 0.01).

**Table 5:**
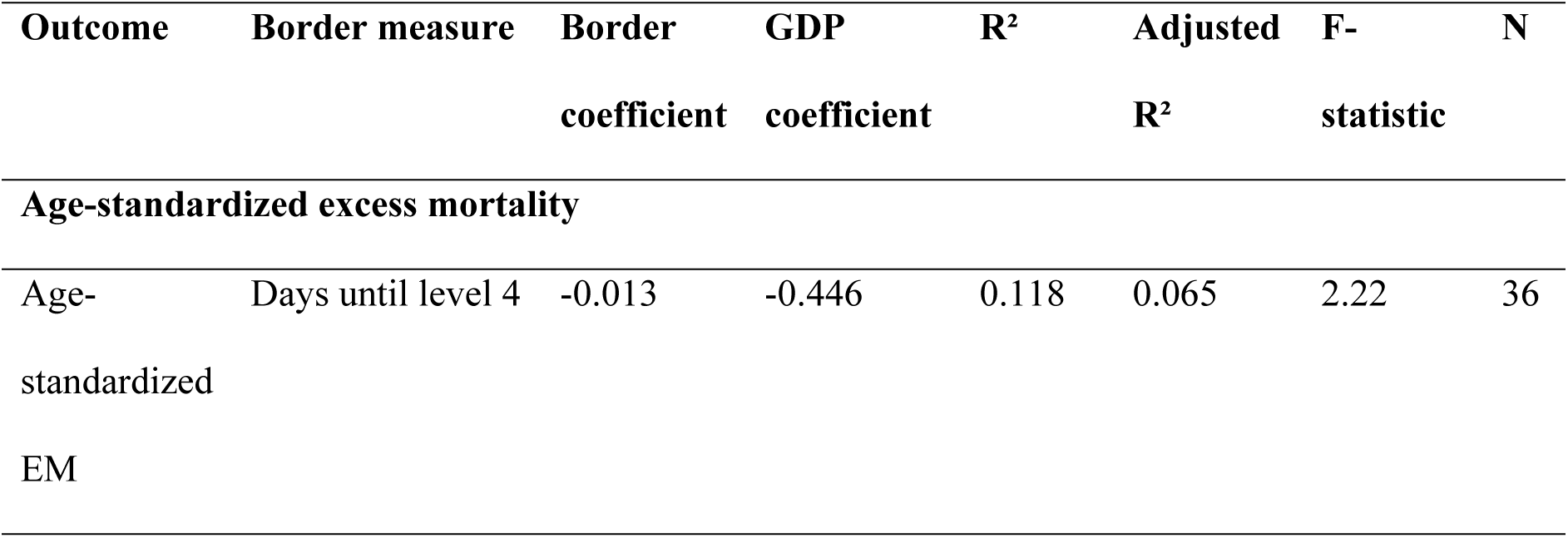
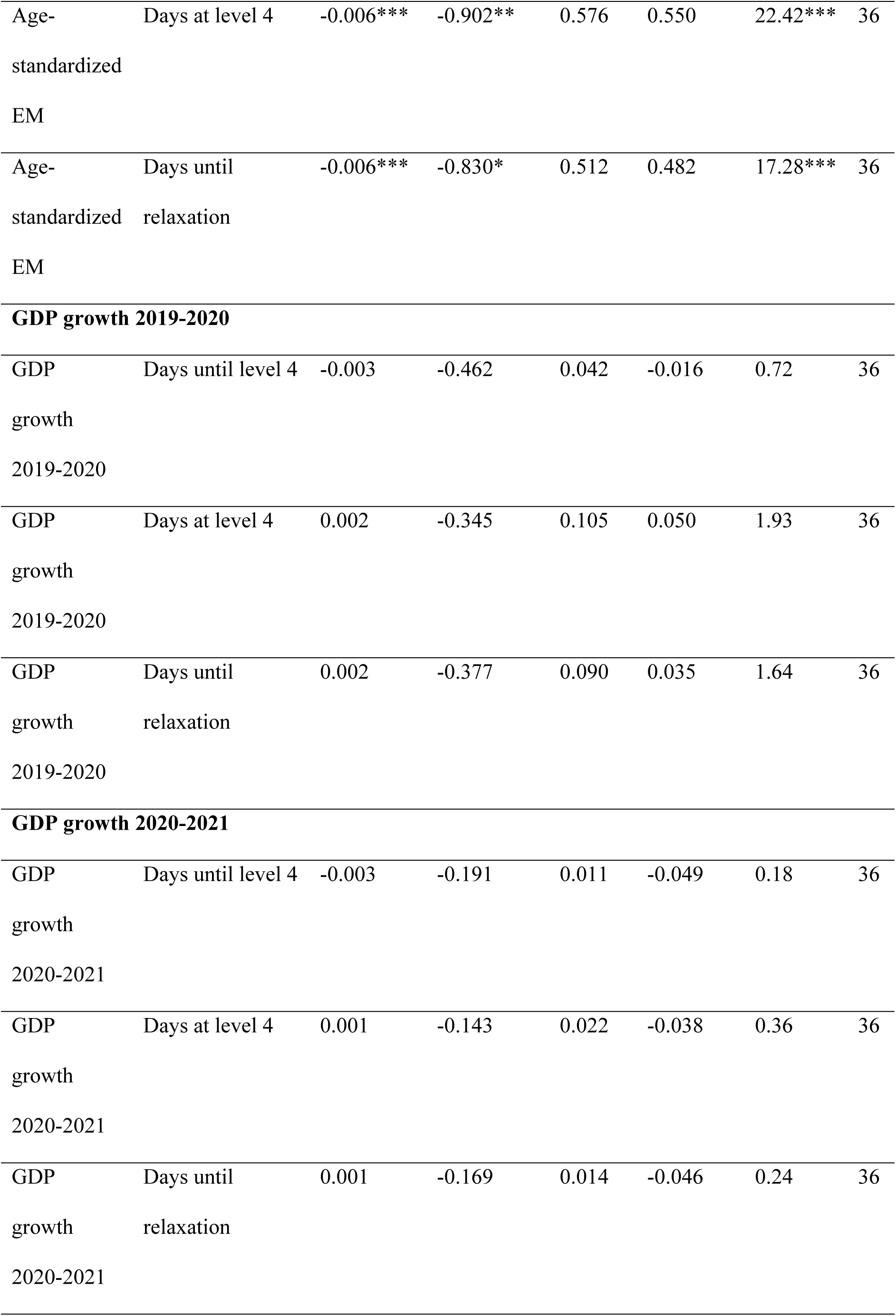
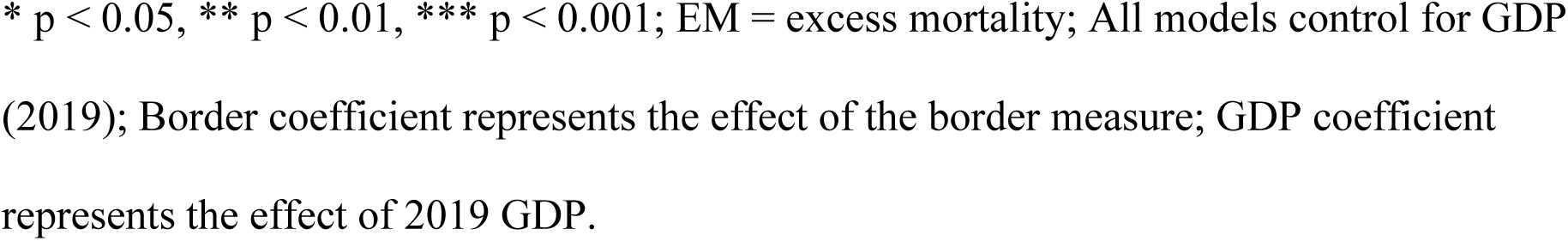
Multivariable regression analysis for island jurisdictions comparing border restrictions and outcomes (excess mortality and GDP growth)

| Outcome | Border measure | Border<br>coefficient | GDP<br>coefficient | $R^2$ | Adjusted<br>$R^2$ | F-statistic | N |
| --- | --- | --- | --- | --- | --- | --- | --- |
| **Age-standardized excess mortality** |  |  |  |  |  |  |  |
| Age-standardized<br>EM | Days until level 4 | -0.013 | -0.446 | 0.118 | 0.065 | 2.22 | 36 |
| Age-standardized<br>EM | Days at level 4 | -0.006*** | -0.902** | 0.576 | 0.550 | 22.42*** | 36 |
| Age-standardized<br>EM | Days until<br>relaxation | -0.006*** | -0.830* | 0.512 | 0.482 | 17.28*** | 36 |
| **GDP growth 2019-2020** |  |  |  |  |  |  |  |
| GDP<br>growth<br>2019-2020 | Days until level 4 | -0.003 | -0.462 | 0.042 | -0.016 | 0.72 | 36 |
| GDP<br>growth<br>2019-2020 | Days at level 4 | 0.002 | -0.345 | 0.105 | 0.050 | 1.93 | 36 |
| GDP<br>growth<br>2019-2020 | Days until<br>relaxation | 0.002 | -0.377 | 0.090 | 0.035 | 1.64 | 36 |
| **GDP growth 2020-2021** |  |  |  |  |  |  |  |
| GDP<br>growth<br>2020-2021 | Days until level 4 | -0.003 | -0.191 | 0.011 | -0.049 | 0.18 | 36 |
| GDP<br>growth<br>2020-2021 | Days at level 4 | 0.001 | -0.143 | 0.022 | -0.038 | 0.36 | 36 |
| GDP<br>growth<br>2020-2021 | Days until<br>relaxation | 0.001 | -0.169 | 0.014 | -0.046 | 0.24 | 36 |

\* $p < 0.05$ , \*\* $p < 0.01$ , \*\*\* $p < 0.001$ ; EM = excess mortality; All models control for GDP (2019); Border coefficient represents the effect of the border measure; GDP coefficient represents the effect of 2019 GDP.

This model explained approximately 58% of the variance in mortality outcomes, which is substantial for a parsimonious model with only two predictors (border restriction duration and GDP per capita). While both variables made meaningful contributions, the higher statistical significance of the border measure duration (p < 0.001 versus p < 0.01 for GDP) suggests it was the more robust predictor. Notably, the models examining economic outcomes (GDP growth 2019–2020 and 2020–2021) showed no significant associations with any border measures, indicating that the mortality benefits of sustained border restrictions in islands were not offset by detectable macroeconomic costs in these jurisdictions during our study period.

However, when controlling for government corruption in a subsample of 19 island jurisdictions with available data, we observed a marked shift in results (Table 6). While the overall model fit improved substantially (R² = 0.714–0.731), the effect of days at level 4 border restrictions on lower mortality was no longer statistically significant (β = –0.002, p > 0.05). Instead, lower government corruption emerged as a significant predictor of lower age-standardized excess mortality (β = 0.697–0.843, p < 0.05). This finding suggests that the apparent protective effect of level ‘4’ border restrictions may be partially confounded by governance quality (at least in this subsample of islands), with better-governed jurisdictions implementing more effective public health measures overall.

**Table 6:**
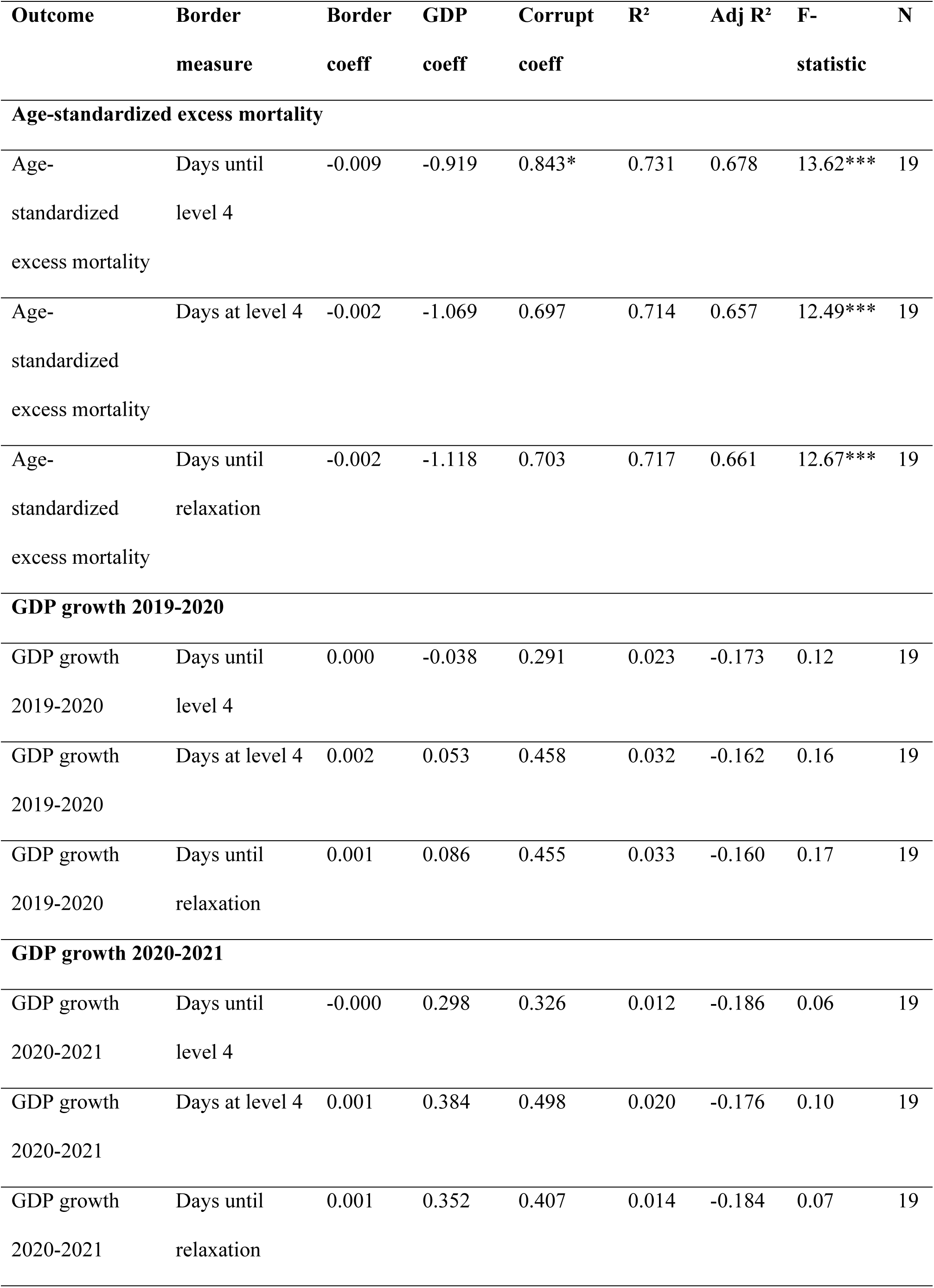
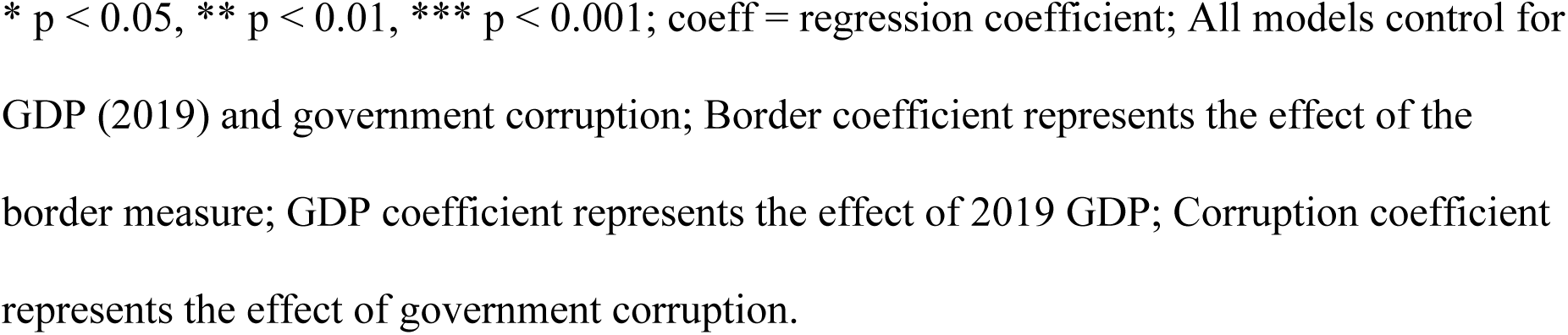
Multivariable regression analysis for island jurisdictions comparing border restrictions and outcomes (excess mortality and GDP growth) and including government corruption index as a control variable (n = 19 subsample of those islands with data)

| **Outcome** | **Border**<br>**measure** | **Border**<br>**coeff** | **GDP**<br>**coeff** | **Corrupt**<br>**coeff** | **R<sup>2</sup>** | **Adj R<sup>2</sup>** | **F-**<br>**statistic** | **N** |
| --- | --- | --- | --- | --- | --- | --- | --- | --- |
| **Age-standardized excess mortality** |  |  |  |  |  |  |  |  |
| Age-standardized<br>excess mortality | Days until<br>level 4 | -0.009 | -0.919 | 0.843* | 0.731 | 0.678 | 13.62*** | 19 |
| Age-standardized<br>excess mortality | Days at level 4 | -0.002 | -1.069 | 0.697 | 0.714 | 0.657 | 12.49*** | 19 |
| Age-standardized<br>excess mortality | Days until<br>relaxation | -0.002 | -1.118 | 0.703 | 0.717 | 0.661 | 12.67*** | 19 |
| **GDP growth 2019-2020** |  |  |  |  |  |  |  |  |
| GDP growth<br>2019-2020 | Days until<br>level 4 | 0.000 | -0.038 | 0.291 | 0.023 | -0.173 | 0.12 | 19 |
| GDP growth<br>2019-2020 | Days at level 4 | 0.002 | 0.053 | 0.458 | 0.032 | -0.162 | 0.16 | 19 |
| GDP growth<br>2019-2020 | Days until<br>relaxation | 0.001 | 0.086 | 0.455 | 0.033 | -0.160 | 0.17 | 19 |
| **GDP growth 2020-2021** |  |  |  |  |  |  |  |  |
| GDP growth<br>2020-2021 | Days until<br>level 4 | -0.000 | 0.298 | 0.326 | 0.012 | -0.186 | 0.06 | 19 |
| GDP growth<br>2020-2021 | Days at level 4 | 0.001 | 0.384 | 0.498 | 0.020 | -0.176 | 0.10 | 19 |
| GDP growth<br>2020-2021 | Days until<br>relaxation | 0.001 | 0.352 | 0.407 | 0.014 | -0.184 | 0.07 | 19 |

\* $p < 0.05$ , \*\* $p < 0.01$ , \*\*\* $p < 0.001$ ; coeff = regression coefficient; All models control for GDP (2019) and government corruption; Border coefficient represents the effect of the border measure; GDP coefficient represents the effect of 2019 GDP; Corruption coefficient represents the effect of government corruption.

A stratified analysis (islands with corruption data [n = 19] vs those without such data [n = 17]) shows that both groups demonstrated similar relationships between duration of level 4 border measures and mortality (β = –0.004, p < 0.05; β = –0.006, p < 0.001, respectively). This finding suggests that selection bias in terms of which countries report corruption data is not driving the original findings. Consistent with most of our primary analysis, we found no significant associations between any border measures and economic outcomes when controlling for corruption.

### Description of jurisdictions with negative excess mortality

Of the seven jurisdictions with negative age-standardized excess mortality during 2020–2021 (Table 7), six were islands (Mongolia being the exception). Four of the seven implemented ‘level 4’ border closure and all used at least some facility quarantine (Taiwan used both types of quarantine and did high quality home quarantine with “digital fencing”). The earliest end of cumulative negative excess mortality was in November 2020 in Iceland (which had incomplete border closure and mainly home quarantine). The mid-year for such a change was 2022 (for Barbados, Japan and Taiwan), and the last year was 2023 (for New Zealand, with Antigua and Barbuda still at negative excess mortality at the end of 2023). This group of jurisdictions included two of the five explicit exclusion/elimination jurisdictions (New Zealand and Taiwan).

## Discussion

### Main findings and interpretation

Our analysis revealed several key patterns regarding the relationship between Covid-19 control strategies, border restrictions, and health outcomes. The five jurisdictions implementing explicit exclusion/elimination strategies demonstrated the lowest mean age-standardized excess mortality, consistent with our first hypothesis. These jurisdictions also maintained the longest median duration of level 4 border restrictions, followed by island jurisdictions more generally, and then non-island jurisdictions.

Our findings confirm previous research [7], showing that island jurisdictions experienced substantially lower excess mortality than non-island jurisdictions, with our results suggesting that relatively stringent border controls may have been a contributing mechanism. However, we observed that some jurisdictions (n = 7, of which six were islands, with high mean GDP per capita) achieved negative excess mortality. Several did so without implementing an explicit exclusion/elimination strategy (Table 7), although they all employed quarantine, regional travel bubbles, and/or level ‘3’ or ‘4’ border restrictions. These finding are consistent with our second hypothesis that effective implementation of stringent border restrictions was associated with lower excess mortality. We note that the absence of any islands in the Oceania region that did not enact ‘level 4’ border restrictions could obscure important relationships between those islands that did and did not implement stringent border controls.

Our analysis reveals a complex relationship between border control measures and pandemic mortality outcomes in island jurisdictions. Initial correlation results and regression models controlling for GDP per capita, demonstrated that longer border restrictions were significantly associated with reduced age-standardized excess mortality (β=-0.006, p<0.001 for days at level 4), suggesting strong protective effects. However, when controlling for government corruption (a methodological improvement that did come at a substantial cost to sample size), this relationship weakened considerably (β=-0.002, p>0.05), while lower government corruption itself emerged as a significant predictor of lower mortality outcomes (β=0.697, p<0.05). Importantly, a stratified analysis comparing jurisdictions with and without corruption data showed consistent negative associations between border measures and mortality in both islands that reported government corruption and those that did not (β=-0.004, p<0.05 and β=-0.006, p<0.001, respectively), indicating the fundamental relationship appears robust to selection bias.

These findings suggest that while relatively stringent border controls likely contributed to reduced pandemic mortality, their effectiveness cannot be fully separated from broader governance factors. The attenuation of border control effects when accounting for corruption indicates these variables share variance in explaining mortality outcomes, reflecting how effective governance may enhance implementation of public health measures. This nuanced picture aligns with emerging pandemic literature highlighting the multifaceted nature of successful responses, where specific interventions operate within broader systems of governance. Future research should continue to explore these complex interactions, as pandemic preparedness requires both specific policy interventions and the governance capacity to implement them effectively.

Regarding economic impacts, neither the original nor the corruption-controlled models revealed consistent relationships between border measures and GDP growth in general. Both sets of models exhibited weak explanatory power, with the corruption-controlled GDP growth models showing negative adjusted R² values that suggest overfitting. This pattern indicates that factors beyond those captured in our models likely played more important roles in determining economic performance during the pandemic period. An important conclusion is that border restrictions in a pandemic may not significantly harm economies.

While this analysis found a strong association between use of a disease exclusion/elimination strategy and low excess mortality, it can only give us limited insights into the mechanism. In the early pandemic phase effective border measures are likely to have reduced mortality directly through preventing outbreaks in fully susceptible populations; genomic data from New Zealand show multiple introductions of the virus prior to implementation of border controls in 2020 [39], and a correspondingly high rate of border introductions in 2022 when border quarantine controls were lifted [40]. The delayed introduction of the virus allowed these jurisdictions to deliver vaccine to their populations and develop improved clinical management before widespread community transmission occurred. Vaccine was only widely available in 2021 which might explain why duration of border restrictions correlated with reduced excess mortality in islands. Some of the benefits of elimination may only become apparent in subsequent years (eg, 2022–2023) when Covid-19, particularly Omicron variants, began circulating widely even in previously protected populations. These additional benefits could include protection against long Covid where the risk is lower for people vaccinated prior to infection [41].

Our metric for speed of border restrictions (days from January 1, 2020, until level 4 implementation), while providing insights also highlighted methodological challenges. For example, New Zealand and Taiwan both implemented level ‘4’ border restrictions at the same time (Table 7), but their response times relative to their first detected cases differed substantially (20 days for New Zealand versus 58 days for Taiwan). Conceptually, rapid border closure following the first detected case aligns closely with an exclusion/elimination approach. Ideally, border controls would be implemented before such cases arrived. These nuances, including time spent with less stringent border controls (level ‘3’, etc) warrant further investigation in future research.

**Table 7:**
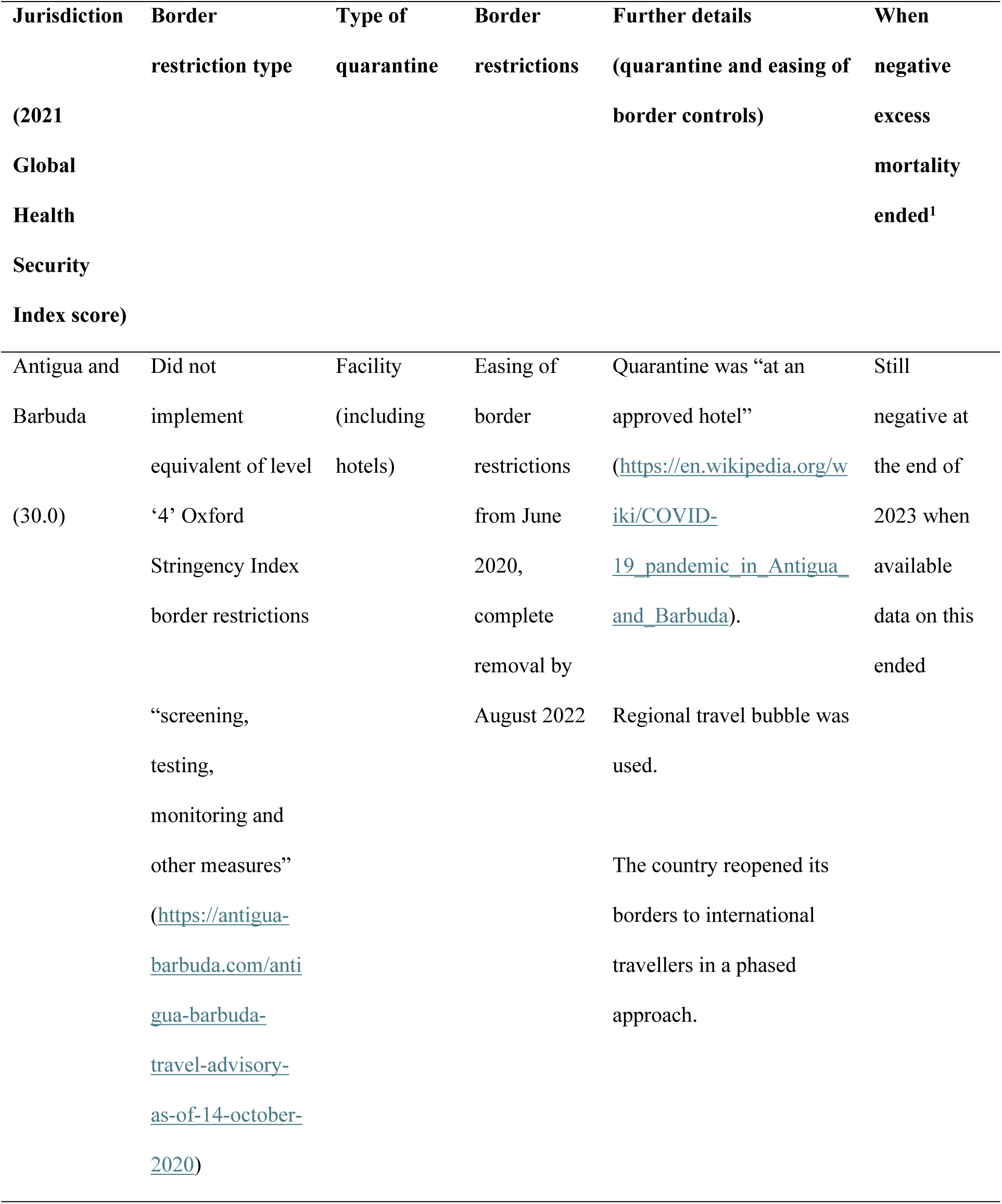
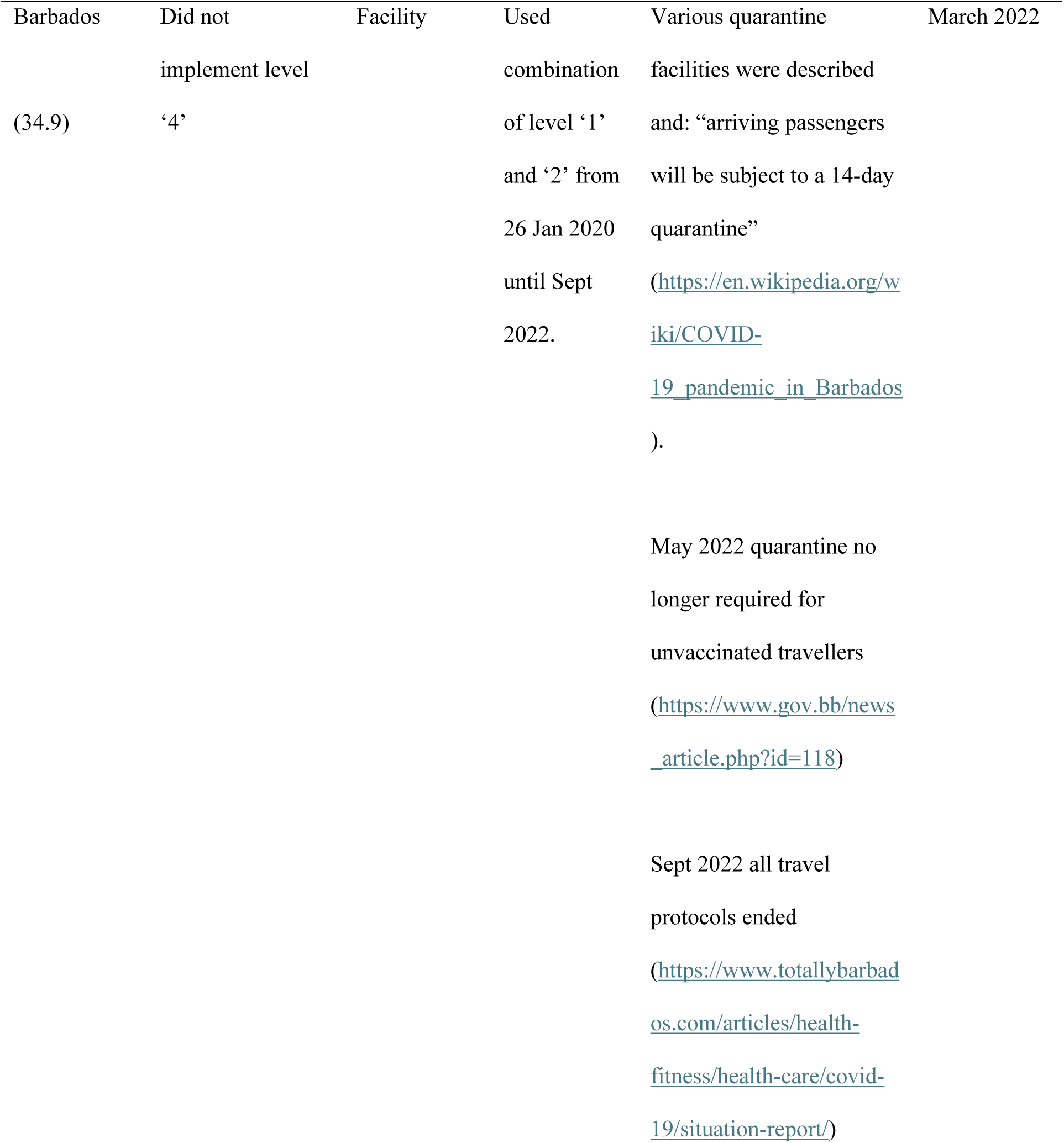
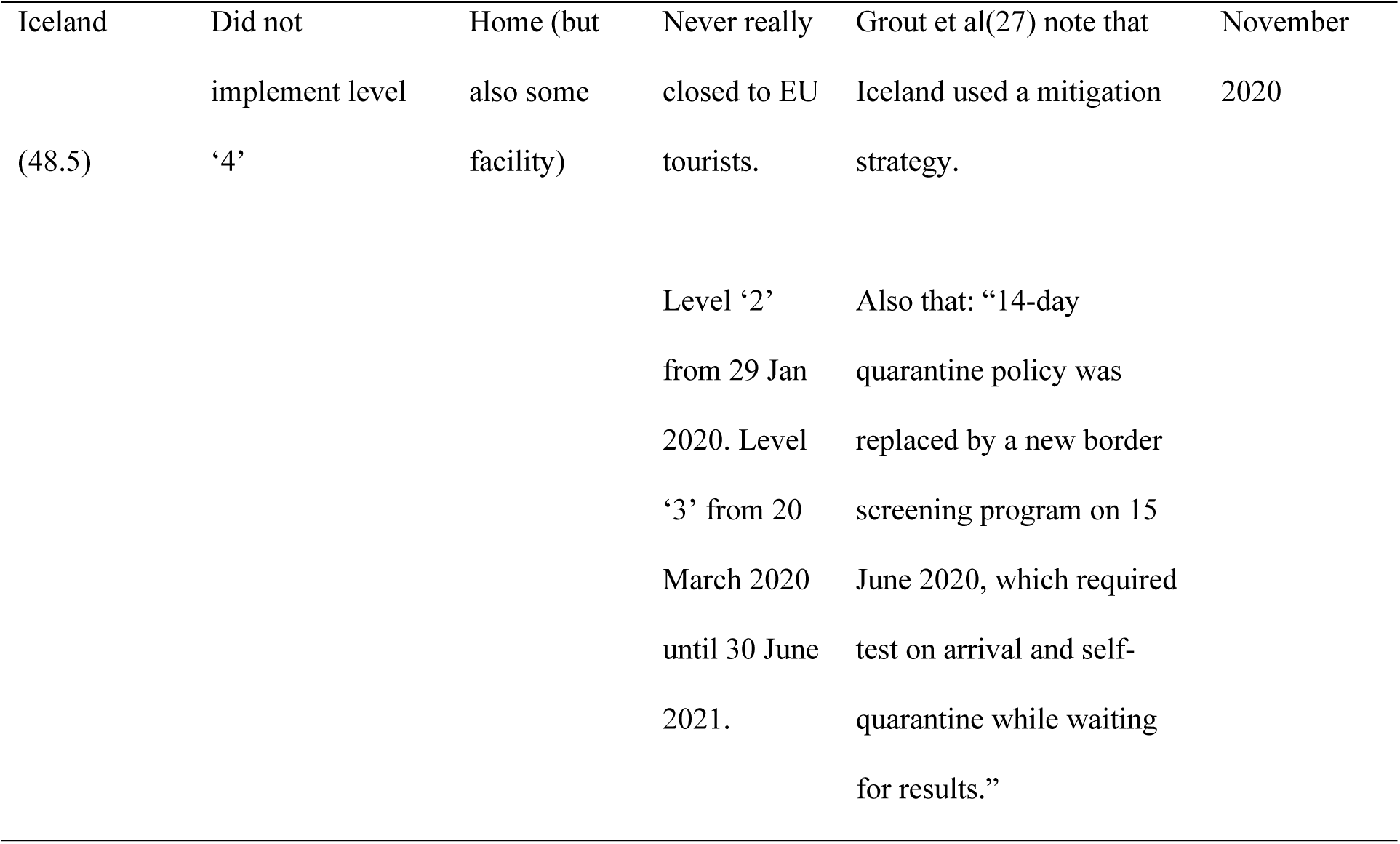
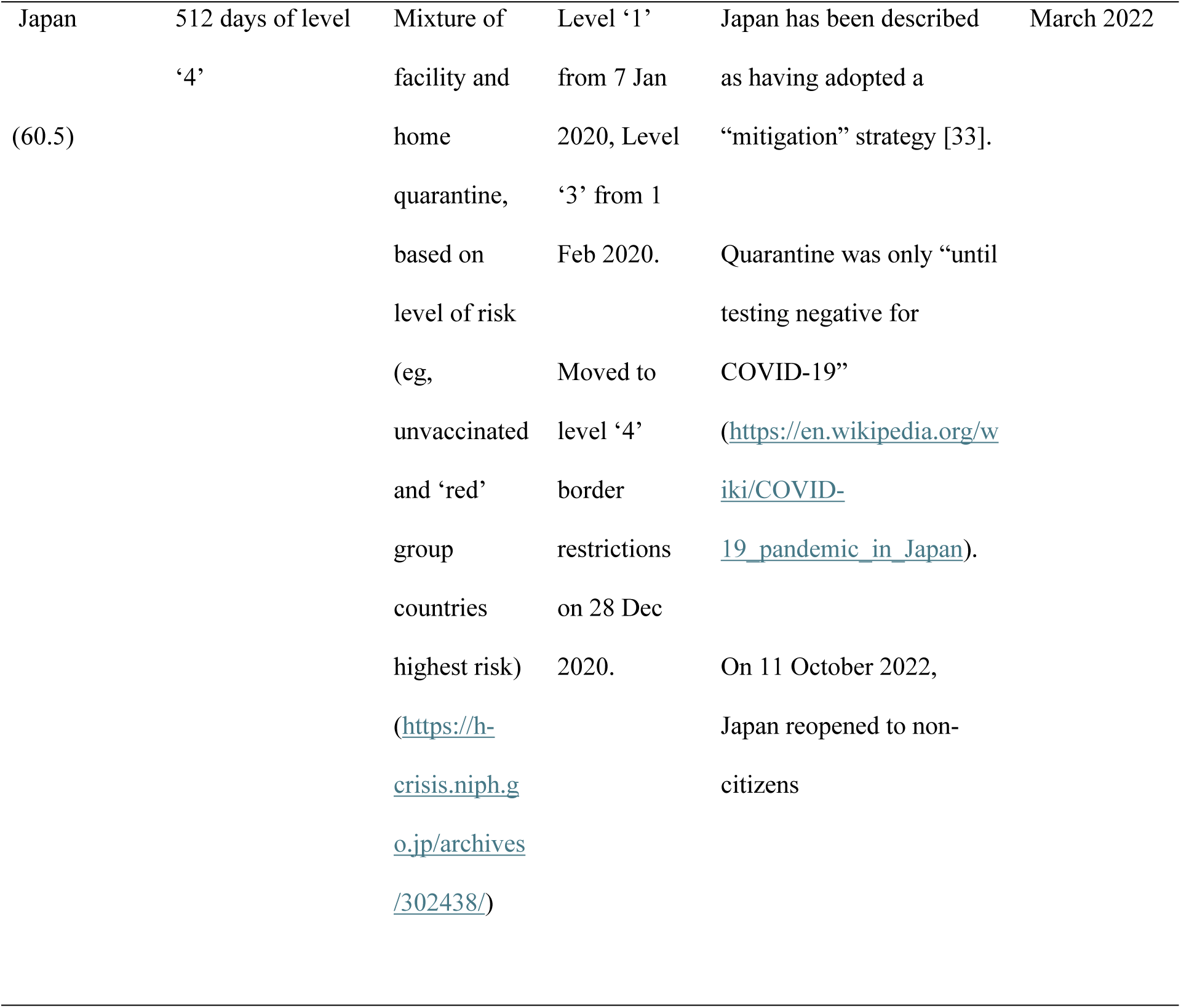
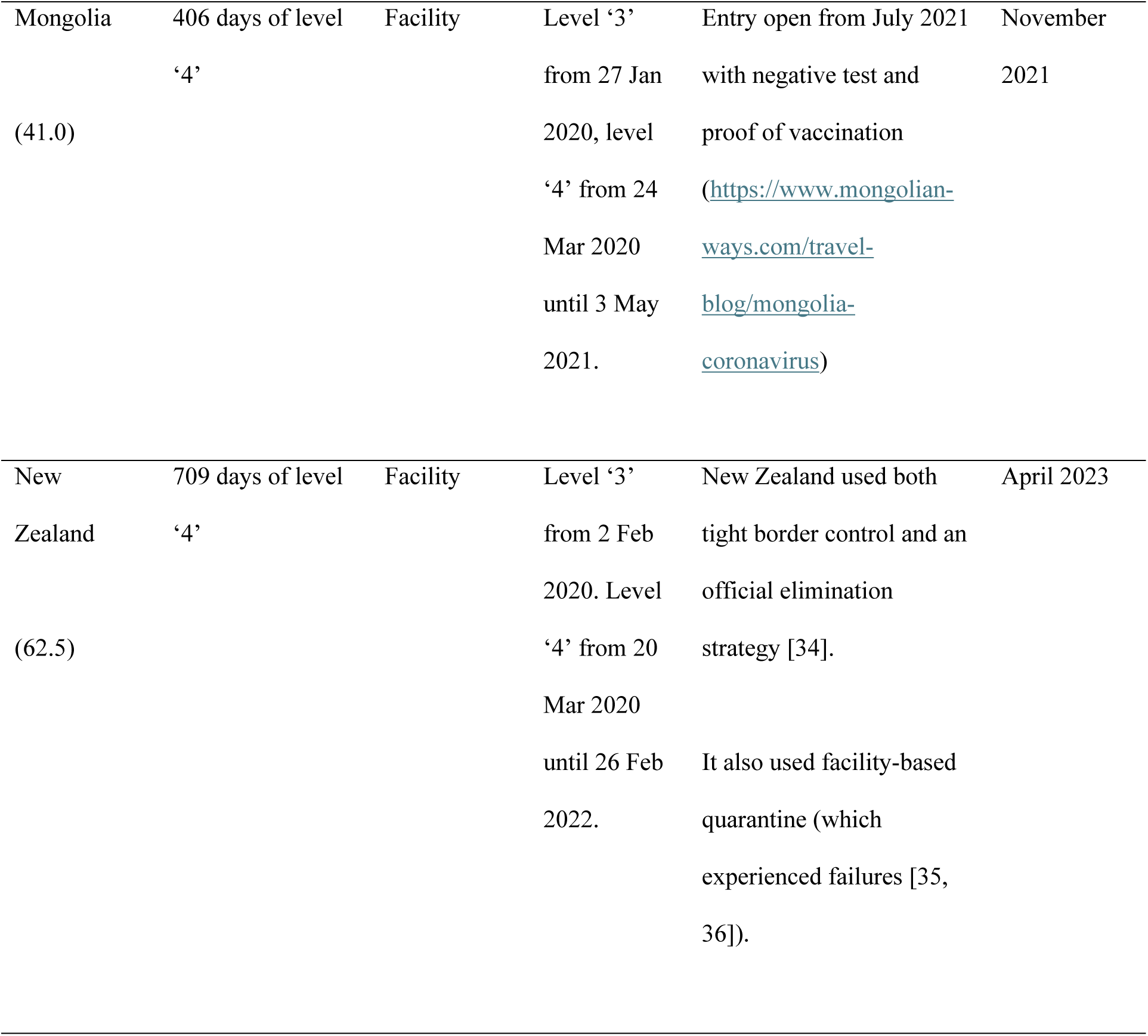
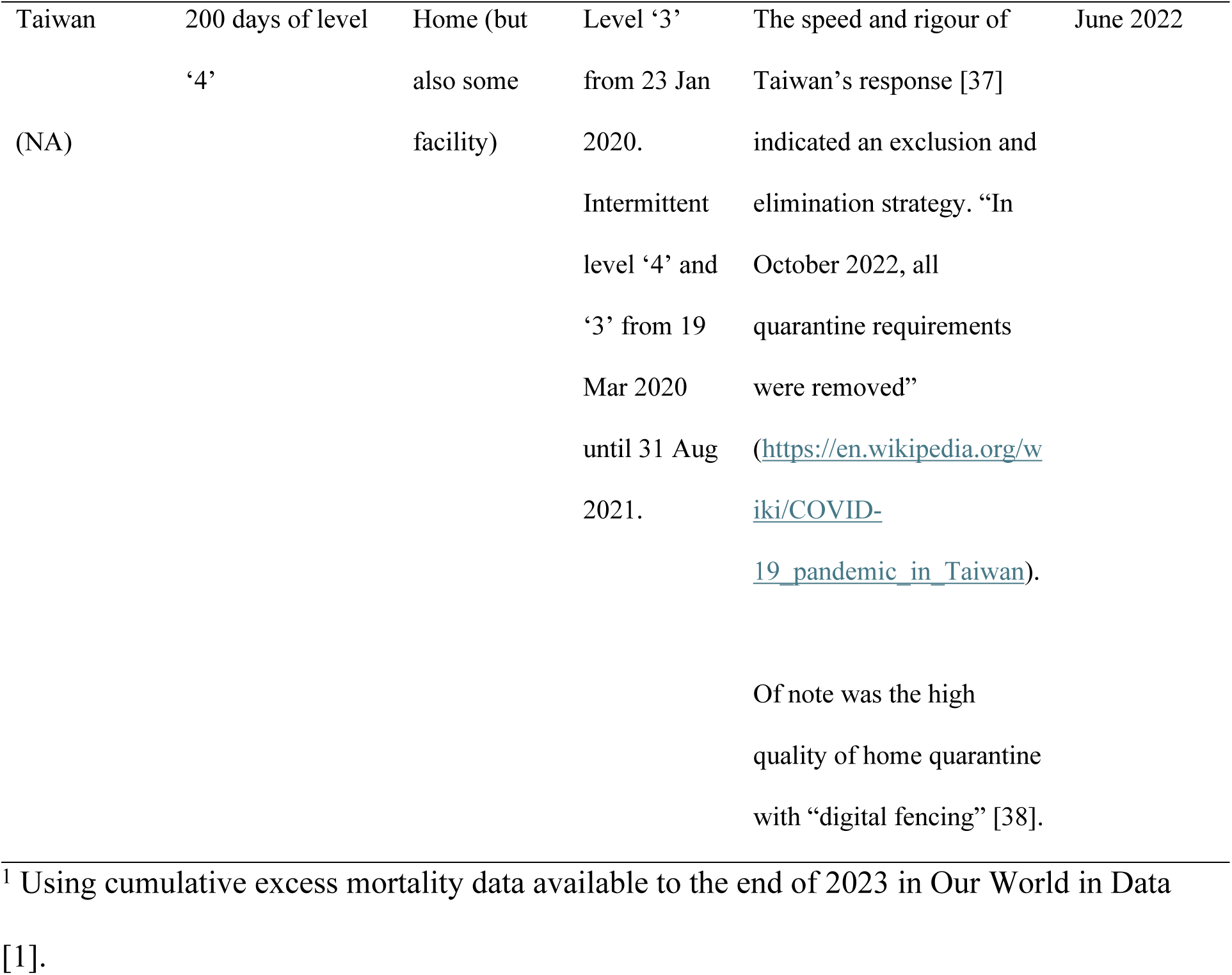
Case studies of jurisdictions with age-standardized negative excess mortality during the period 2020 to 2021.

### Study strengths and limitations

Our study has several notable strengths, including the use of a comprehensive global dataset covering 193 jurisdictions with age-standardized excess mortality data. We employed a novel analytical approach examining the timing of “complete” border restrictions. Our multivariable analyses controlled for key confounding factors including GDP per capita and island status, while our exploration of government corruption as an additional control variable provided important insights into the role of governance quality during pandemic response.

However, several limitations should be acknowledged. Our analysis focused solely on sovereign jurisdictions, excluding sub-national entities that implemented successful exclusion/elimination strategies (eg, Hong Kong [42], as did various Canadian Provinces [14]). Mean age-standardized excess mortality for the excluded non-sovereign islands (30.2/100,000) was lower than other islands, so this is likely a conservative exclusion, ie, making it harder to find differences in excess mortality between islands and non-islands.

The most stringent level ‘4’ of border closures may not accurately reflect actual enforcement, with potential discrepancies between reported policies and implementation, including possible circumvention via unofficial border crossings.

Additionally, our analysis including a government corruption metric was limited to jurisdictions with available data, potentially introducing selection bias and reducing statistical power. The Global Burden of Disease Study excess mortality estimates have inherent methodological limitations (eg, for both mortality and demographic variables [25]). Furthermore, our analysis does not capture longer-term outcomes through subsequent years (eg, 2022–2024). Important confounding factors such as trust in government, population health metrics (eg, burden of chronic illness), and Covid-19 vaccination coverage were also not included in our adjustments. Finally, economic impact assessments were complicated by the potential disproportionate effect on tourism-dependent economies and the potential economic benefits experienced by countries exporting information technology equipment and pharmaceutical and medical technology products during the pandemic.

This analysis is entirely based on the Covid-19 pandemic, a virus spread via the respiratory route and with specific, evolving infectious diseases dynamics. Consequently, our findings may have reduced generalisability to pandemics with very different characteristics, though the most probable future severe pandemic threats such as influenza share many of the same features [43]. It is notable that respiratory control measures used during the emergency phase of the Covid-19 pandemic eliminated transmission of a number of non-pandemic pathogens [44], including potential extinction of influenza B/Yamagata virus [45].

### Implications for policy and research

Our findings suggest that while exclusion/elimination strategies and relatively stringent border controls may be associated with better health outcomes, particularly for island jurisdictions, the effectiveness of these approaches was likely partially mediated by broader governance factors including institutional quality and corruption control. This conclusion highlights the importance of considering not just what policies are implemented, but the quality and context of their implementation.

For future pandemic preparedness, our research suggests that border control strategies may be particularly effective for island jurisdictions, but their success likely depends on broader governance capabilities, resources, and institutional integrity. Simply implementing strict border measures without addressing underlying governance issues may not achieve optimal outcomes. Such measures could include the use of predetermined “green zones” (travel bubbles) and improved measures to prevent introduction of the infection by incoming travellers, enabling movement of people but not pathogens across borders.

Future research could attempt to better categorize pandemic response strategies of countries, notably distinguishing all of those that pursued exclusion/elimination and the levels of control they achieved. It could further explore the complex relationships between governance quality, policy implementation, and pandemic outcomes using more comprehensive datasets that include measures of corruption and other social and governance metrics previously found to predict pandemic outcomes [6], for a wider range of jurisdictions. We were unable to meaningfully test many of these questions due to poor data coverage for islands.

Additionally, examining longer-term outcomes beyond our study period could provide valuable insights into the enduring impacts of different strategic approaches to pandemic control. Research could also assess how non-island countries such as Mongolia were also able to use and in some cases sustain exclusion/elimination strategies for prolonged periods without the border control advantages of island countries.

Global standard setting organisations such as the World Health Organization have an important role in providing a consistent strategic framework to support pandemic prevention, preparedness and response. As part of this role they should investigate the use of standard terminology for pandemic response strategies as a key organising concept. Such a framework could also be integrated into the International Health Regulations and the process of responding to public health emergencies of international concern [46].

## Conclusions

Our analysis of 193 jurisdictions provides insights into Covid-19 pandemic control strategies during 2020–2021. Jurisdictions implementing explicit exclusion/elimination strategies demonstrated substantially lower excess mortality than others, along with island nations with stringent border restrictions. While extended strict border control measures were associated with reduced mortality in island jurisdictions, this relationship weakened when controlling for government corruption, suggesting governance quality as a crucial mediator of policy effectiveness. Importantly, we found no consistent significant associations between border measures and GDP growth, challenging assumptions about inevitable health-economy trade-offs with strict border controls.

These findings suggest that exclusion/elimination may be the optimal pandemic control strategy where it can be achieved. Circumstances supporting this response include island jurisdictions and strong governance capabilities. Future pandemic preparedness should therefore consider both geographical context and governance quality when designing response strategies. Governance quality is modifiable. While geography is fixed, there are ways of achieving comparable border control attributes of islands with advanced planning.

Such measures could include “green zones” and improved measures to prevent introduction of the infection by travellers. As the world prepares for future emerging infectious disease threats, these insights can inform more proactive approaches to pandemic prevention, preparedness and response.

## Acknowledgements

Prof Austin Schumacher from the GBD Collaboration for data sharing.

## Data availability

The datasets used are available from the authors on request (GBD Study data to be requested from GBD Study authors).

